# FREQUENCY SPECIFICITY OF NARROWBAND CHIRP AND 2-1-2 STIMULI: SPECTRAL ANALYSES

**DOI:** 10.1101/2024.11.09.24317036

**Authors:** Ronald Adjekum, Susan A. Small, Sylvia Chan, David R. Stapells

## Abstract

**Objective:** The current study examined the frequency specificity of NB chirps by comparing the spectral characteristics of 500-, 1000-, 2000- and 4000-Hz NB CE-Chirp^®^ LS stimuli with those of 2-1-2 tones.

**Design:** Spectral characteristics including the centre frequency, bandwidth, and stimulus energy changes after stopband filtering were compared. The bandwidth was computed as the difference between the upper and lower frequencies at -20 dB (& -3 dB) cutoff points of the main lobe; the centre frequency was determined as the geometric mean of the upper and lower frequencies at the -20 dB (& -3 dB) cutoff points.

**Results:** At 100 dB peSPL, the bandwidths of the 500-, 1000-, and 2000-Hz NB CE-Chirp^®^ LS acoustic spectra were 1.7-2.5 times wider than the acoustic spectra for the 2-1-2 tones; the 4000-Hz NB CE-Chirp^®^ LS bandwidths were 1.4-1.6 times wider than those of the 2-1-2 tones. The energy of NB CE-Chirp® LS stimuli was concentrated within ±0.75 octave of the centre frequency, compared to ±0.5 octave for 2-1-2 tones.

**Conclusion:** NB CE-Chirp^®^ LS stimuli demonstrated poorer frequency specificity compared with 2-1-2 tones. Further studies are needed to investigate the place specificity of the ABRs to NB CE-Chirp^®^ LS before implementing them clinically.

## Introduction

Physiological assessment tools, such as the auditory brainstem response (ABR), are used to evaluate hearing sensitivity in populations for which behavioural testing is a challenge, especially for young infants who are at risk for hearing loss. Estimating frequency-specific thresholds and assessing the neural integrity of the auditory pathways are essential for implementing early intervention for hearing loss before six months of age (The Joint Committee on Infant Hearing, 2019). The type of stimulus used in testing is critical in achieving these goals. To estimate hearing threshold, the aim is to use a stimulus that elicits a response with as large an amplitude as possible without losing frequency specificity.

Over 30 years ago, Davis and colleagues recommended tonal stimuli with linear rise/fall times of 2 cycles each and a plateau of 1 cycle (“2-1-2” tones) as the best compromise between acoustic frequency specificity and stimulus duration (Davis, 1976). Because ABR wave V amplitudes show large decreases when stimulus rise times exceed 5 ms, Stapells and Picton (1981) concluded that a rise/fall time of 5 ms (giving a total duration of 10 ms) represented a good compromise between frequency specificity and response recognizability. Specifying stimuli such that they contain a constant number of cycles (rather than constant milliseconds), however, has been favoured as doing so results in similar stimulus frequency specificity regardless of stimulus frequency. Although some researchers have recommended the use of shorter tonal stimuli (e.g., Gorga & Thornton, 1989; McCreery et al., 2015), many studies have demonstrated that ABR thresholds to 2-1-2 tones (and other brief tones with similar duration, such as 5-cycle Blackman-windowed) provided reasonable estimates of the pure-tone audiograms in individuals with normal or impaired hearing (for review, see: Small & Stapells, 2017).

More recently, “chirp” stimuli have been of interest to elicit ABRs both for hearing screening purposes using broadband chirps and for threshold estimation using octave-band (“narrowband”) chirps. This interest is largely due to studies indicating chirps (both broadband and narrowband) usually (but not always) elicit larger ABR amplitudes compared to more-standard stimuli (i.e., clicks and 2-1-2 stimuli) (Bell et al., 2002; Cobb & Stuart, 2016; Dzulkarnain et al., 2018; Ferm & Lightfoot, 2015; Rodrigues et al., 2013). Larger response amplitudes should improve the signal-to-noise ratio (SNR) of the ABR and help to reduce testing time (Bell et al., 2002; Burkard & Don, 2007; Ferm & Lightfoot, 2015)

When broadband clicks are used to elicit an ABR, due to the stiffness properties of the cochlear partition, the low-frequency components reach their place of maximum displacement farther along the cochlear partition, and thus later, than the higher frequency components. This produces temporal delays in the brainstem response from different cochlear (and thus frequency) regions, in essence producing multiple waves V occurring at differing times from each cochlear region (Don & Eggermont, 1978). The timing differences partially cancel the scalp-recorded ABR for some frequency components (Burkard & Don, 2007). In contrast, a broadband chirp stimulus rising in frequency with the temporal spacing of the frequency components of the stimulus adjusted such that maximum neural firing across the basilar membrane occurs simultaneously, resulting in less cancellation and larger ABR amplitudes.

Rising frequency chirp stimuli were initially developed using cochlear delay models based on the linear characteristics of the cochlea (Dau et al., 2000). Subsequent studies focussed on modelling chirps that most efficiently elicited the ABR (Elberling et al., 2010). The aim was to elicit greater discharge synchronicity and, consequently, larger ABR amplitudes than standard click stimuli (Dau et al., 2000; Elberling & Don, 2010; Kristensen & Elberling, 2012). Indeed, studies in adult humans have confirmed that ABRs to broadband chirps are usually significantly larger than to clicks (Dau et al., 2000; Elberling et al., 2010; Kristensen & Elberling, 2012; Wegner & Dau, 2002). Depending on the type of chirp stimulus (or delay model), this advantage may not exist at high (e.g., 80 dB nHL) stimulus levels (e.g., Kristensen & Elberling, 2012). For example, at 80 dB nHL, Kristensen and Elberling (2012) showed that Wave V amplitude to click was similar to that of a level-independent chirp (CE-Chirp^®^) but significantly smaller than the responses to a level-specific chirp (CE-Chirp^®^ LS). Results for the broadband CE-Chirp^®^ and CE-Chirp^®^ LS designed by Elberling and colleagues are the most commonly reported in the ABR and auditory steady-state response literature and were based on derived-band ABR latencies (Elberling et al., 2007; Elberling & Don, 2010).

Recently, there has been much interest in using narrowband chirps to estimate frequency-specific ABR thresholds. Narrowband chirps (NBchirps) have been generated by decomposing the broadband CE-Chirp^®^ into four octave-band chirps (centre frequencies: 500, 1000, 2000 & 4000 Hz) (Elberling et al., 2007) and are available in clinical equipment (e.g., Interacoustics Eclipse). The NB CE-Chirp^®^ stimuli were designed to increase the synchronous activities of auditory neural fibres, which would potentially elicit larger response amplitudes compared to ABRs to more-standard stimuli such as 2-1-2 tones. This is despite the earlier conclusion by Wegner and Dau (2002) that one-octave-wide regions may not be sufficiently wide enough to produce an amplitude advantage over 2-1-2 stimuli. That is, stimulation of the NBchirp restricted to a one-octave-wide basilar membrane region would be too small for the optimized timing of a NBchirp to make a significant difference in amplitude. This may partially explain why the amplitude advantage of NB CE-Chirp^®^ is not consistently present (e.g., Cobb & Stuart, 2016; Dzulkarnain et al., 2018; Ferm & Lightfoot, 2015; Rodrigues et al., 2013)

Inconsistent ABR amplitude results for NB CE-Chirp^®^ stimuli at different intensities are not surprising as earlier studies of broadband chirp stimuli had noted that the amplitude advantage was not present at higher stimulus levels, presumably due to the spread of excitation along the basilar membrane and superimposition of responses from the low- and high-frequency regions Current NBchirp stimuli are usually described as “octave-band” stimuli (e.g., Elberling & Don, 2010; Gøtsche-Rasmussen et al., 2012). The frequency spectra of NBchirps appear to be wider than 2-1-2 tones, although their bandwidths have not been specifically quantified. A figure by Bell, Allen, and Lutman (2002) showed that the spectra of NBchirps, which are approximately 1-octave-wide at half power, are much wider than those of 2-1-2 tones, which led Bell and colleagues to suggest that NBchirp spectra may be too wide for clinical application.

Cobb and Stuart (2016) also presented acoustic spectra showing that the narrow band CE-Chirp^®^ main lobes were wider than the linear brief tones and suggested the wider spectra of NBchirps may contribute to larger ABR amplitudes. Neither study provided quantitative descriptions of the acoustic characteristics of NBchirps beyond the spectra shown in the figures.

The importance of frequency specificity of stimuli used to elicit the ABR has long been an issue, often heated, in the field of Audiology (e.g., Davis, 1976; Gorga & Thornton, 1989; Laukli, 2014; Laukli et al., 1988; Stapells et al., 1994). The lack of frequency specificity of clicks (and broadband chirps) means they have limited utility for threshold estimation (Burkard & Don, 2007; Davis, 1976; Stapells et al., 1994). In contrast, many feel brief tones (such as 2-1-2 tones) provide acceptable, albeit imperfect, frequency specificity (Davis et al., 1984; McCreery et al., 2015; Small & Stapells, 2017), although not all agree (Burkard & Hecox, 1983; Gorga & Thornton, 1989; Laukli, 2014). In more recent years, there seems less concern around the frequency specificity of the brief-tone stimuli typically used to elicit the ABR (such as 2-1-2 tones), and these are currently used in large-scale early hearing programs (e.g., Bagatto, 2020; Hatton et al., 2022; Te Whatu Ora – Health New Zealand, 2023). The frequency specificity (i.e., spectral content) of brief stimuli such as 2-1-2 tones are well known (e.g., Burkard, 1984; Davis, 1976; Davis et al., 1984; Gorga & Thornton, 1989). In contrast, although gaining in popularity and clinical use, to date issues regarding the frequency specificity of NBchirps have seen little-to-no discussion, with limited comparison of NBchirps and 2-1-2 tones. The objective of the present study is to investigate this issue further by providing a detailed comparison of the spectral features of the NB CE-Chirp^®^ LS and 2-1-2 stimuli at 500, 1000, 2000, and 4000 Hz.

## Methods

### Stimuli and calibration

Stimuli (500, 1000, 2000, and 4000 Hz) were generated by an Interacoustics Eclipse ABR system with research version EPx5 4.4 software. The acoustic waveforms were generated by presenting NB CE-Chirp^®^ LS and 2-1-2 stimuli at 60, 80, and 100 dB pe SPL via ER-3A insert earphones.^1^ The stimulus levels were calibrated and recorded using a GRAS RA0113 2-cc coupler, a 1-inch microphone (Larson Davis Model 2575), and a PRM902 preamplifier with a Larson Davis System 824 sound level meter. The sound level meter output was routed to a PC for analyses by Sigview software (version 3.2), using a 44,100 Hz digitation rate and a 95-ms analysis window. Each recorded stimulus was obtained after averaging 100 time-domain samples. For electrical waveform analyses, which were only carried out on the 100 dB pe SPL stimuli, the stimulus output of the Interacoustics Eclipse ABR system was routed directly to the PC for analyses by the Sigview software, using the same settings in Sigview as for analyses of the acoustic waveforms.

### Spectral analyses

Spectral content for each stimulus was derived from a Fast Fourier transform (FFT) performed using the Sigview software, using a Hamming window and bin size of 4096 points. The frequency specificity of the NB CE-Chirp^®^ LS and 2-1-2 stimuli was determined by analyzing their spectra for their bandwidth (in Hz & octaves), centre frequency (Hz), and peak frequency. The bandwidth for each stimulus was calculated based on upper and lower frequency cutoff points at 3 dB and 20 dB below the peak energy of the stimulus. Although the -3 dB cutoffs are typically used to describe filters and were also used by Elberling and colleagues to describe the bandwidth of NBchirps (Elberling & Don, 2010; Gøtsche-Rasmussen, et al., 2012), the -3-dB bandwidth underestimates the physiological effective bandwidth (i.e., more than the top 3 dB typically contribute to a response)(e.g., Oates & Stapells, 1997; Purdy & Abbas, 2002). We therefore calculated the -20-dB bandwidth as a more realistic estimate of *effective* bandwidth contributing to a response. For the same reason, centre frequencies were determined using the upper and lower frequency cutoff points at -20 dB. The bandwidth was determined by calculating the difference between the upper and lower frequency cutoffs, whereas centre frequencies were measured as the geometric mean (GM) of the same at -3 and -20 dB. The peak frequency for each stimulus was a measure of the frequency with the highest amplitude. A comparison between electrical and acoustic spectra was conducted to analyze the effect of the ER-3A insert phone on the NB CE-Chirp^®^ LS versus 2-1-2 stimuli. Finally, the energy remaining after 0.5-, 1-, 1.5-, 2-, and 3-octave-wide stopband filtering of the stimulus waveforms was computed as a percentage of the stimulus energy before filtering.

## Results

Figure 1 (left) shows the acoustic time-domain waveforms of the NB CE-Chirp^®^ LS and 2-1-2 tones for 500-, 1000-, 2000-, and 4000-Hz stimulus frequencies. The latencies of the maximum amplitudes, relative to the onset of the stimulus, were later for NBchirps compared to 2-1-2 stimuli.^2^ Figure 1 (right) also shows the spectra of the acoustic stimulus waveforms. The main lobes of the NB CE-Chirp^®^ LS acoustic spectra were broader than those of the 2-1-2 tone acoustic spectra. The 4000-Hz NB CE-Chirp^®^ LS acoustic spectra appeared primarily as one relatively wide main lobe (Figure 1, right). The acoustic spectra for 2-1-2 tones showed narrow main lobes with well-defined lobes of sideband energy.

**Figure 1.**
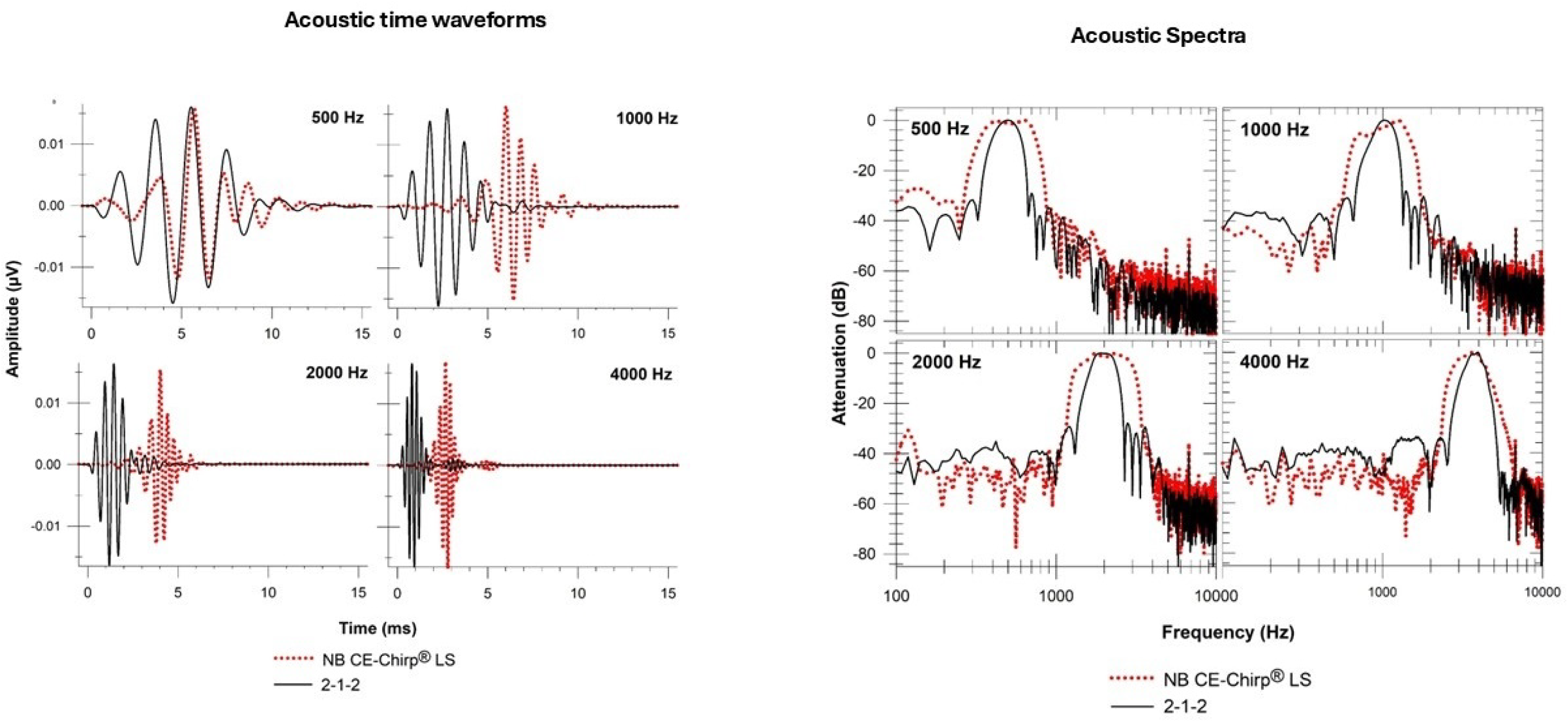
Time-domain waveforms (left) and acoustic spectra (right) for the 500-, 1000-, 2000-, and 4000-Hz NB CE-Chirp® LS and 2-1-2 stimuli. (NB chirps: red dotted lines; 2-1-2 tones: black solid lines.)

Table 1 presents the measures from the acoustic spectra for NB CE-Chirp^®^ LS and 2-1-2 stimuli presented at 60, 80, and 100 dB pe SPL. As is obvious from Table 1, there were no large or consistent differences in the measures between the 60, 80, and 100 dB pe SPL acoustic spectra. This is because, other than a difference in stimulus onset timing, the same delay model was used for all the Eclipse “NB CE-Chirp^®^ LS” stimulus intensities. That is, the NB CE-Chirp^®^ LS stimulus presented by the Eclipse (research version EPx5 4.4) does not change waveforms with increasing stimulus intensity.^3^, the remainder of the Results presentation will focus on the 100 dB pe SPL results.

**Table 1.** Acoustic spectral characteristics for NB CE-Chirp^®^ LS versus 2-1-2 tones at 60, 80, and 100 dB pe SPL.

| Stimulus Type | Intensity (dB peSPL) | Frequency (Hz) |  | GM (Hz) |  | BW (Hz) |  | BW (octaves) |  |
| --- | --- | --- | --- | --- | --- | --- | --- | --- | --- |
|  |  | Nominal | Peak | -3dB | -20dB | -3dB | -20dB | -3dB | -20dB |
| NBCh | 60 | 500 | 635 | 509 | 486 | 335 | 536 | 0.9 | 1.5 |
| 212 TB |  |  | 506 | 496 | 479 | 136 | 290 | 0.4 | 0.9 |
| NBCh | 60 | 1000 | 1238 | 1175 | 1001 | 371 | 1055 | 0.5 | 1.5 |
| 212 TB |  |  | 1034 | 1032 | 963 | 247 | 578 | 0.3 | 0.9 |
| NBCh | 60 | 2000 | 1766 | 2115 | 2012 | 1317 | 2156 | 0.9 | 1.5 |
| 212 TB |  |  | 2035 | 1984 | 1918 | 594 | 1147 | 0.4 | 0.9 |
| NBCh | 60 | 4000 | 3596 | 3521 | 3592 | 1180 | 3017 | 0.5 | 1.2 |
| 212 TB |  |  | 3973 | 3800 | 3695 | 679 | 2033 | 0.3 | 0.8 |
| NBCh | 80 | 500 | 635 | 505 | 485 | 340 | 540 | 1.0 | 1.5 |
| 212 TB |  |  | 495 | 494 | 477 | 139 | 296 | 0.4 | 0.9 |
| NBCh | 80 | 1000 | 1238 | 1159 | 994 | 391 | 1066 | 0.5 | 1.5 |
| 212 TB |  |  | 1023 | 1029 | 963 | 247 | 591 | 0.3 | 0.9 |
| NBCh | 80 | 2000 | 1766 | 2104 | 1995 | 1305 | 2153 | 0.9 | 1.5 |
| 212 TB |  |  | 1895 | 1981 | 1918 | 600 | 1155 | 0.4 | 0.9 |
| NBCh | 80 | 4000 | 3542 | 3499 | 3581 | 1138 | 2903 | 0.5 | 1.2 |
| 212 TB |  |  | 3930 | 3778 | 3701 | 705 | 2051 | 0.3 | 0.8 |
| NBCh | 100 | 500 | 635 | 504 | 484 | 341 | 542 | 1.0 | 1.5 |
| 212 TB |  |  | 495 | 494 | 476 | 139 | 297 | 0.4 | 0.9 |
| NBCh | 100 | 1000 | 1238 | 1143 | 989 | 420 | 1074 | 0.5 | 1.5 |
| 212 TB |  |  | 1023 | 1026 | 959 | 244 | 589 | 0.3 | 0.9 |
| NBCh | 100 | 2000 | 1766 | 2092 | 1995 | 1321 | 2159 | 0.9 | 1.5 |
| 212 TB |  |  | 1895 | 1980 | 1917 | 594 | 1159 | 0.4 | 0.9 |
| NBCh | 100 | 4000 | 3564 | 3502 | 3588 | 1136 | 2869 | 0.5 | 1.1 |
| 212 TB |  |  | 3941 | 3777 | 3705 | 709 | 2045 | 0.3 | 0.8 |

Peak Frequency: Frequency with highest amplitude; GM: Geometric mean; BW: Bandwidth; NBCh: NB CE-Chirp<sup>®</sup> LS; 212 TB: 2-1-2 tone; -20dB: measured at 20 dB below maximum amplitude; -3dB: measured at 3 dB below maximum amplitude

Across all frequencies, peak frequencies (the frequency showing maximum amplitude) for NBchirps were a little lower than the nominal frequencies (approximately -6%), whereas 2-1-2 tones were closer to the nominal frequencies (within 1%). The geometric mean frequencies were very close to the nominal frequencies, especially measured at -3dB. The -3-dB bandwidths for the NB CE-Chirp^®^ LS stimuli were close to one octave in width, as previously indicated by Elberling and colleagues (Elberling & Don, 2010; Gøtsche-Rasmussen, et al., 2012). However, comparing the -3-dB bandwidths between stimuli, the NB CE-Chirp^®^ LS spectra were considerably wider than those of the 2-1-2 tones. For example, NB CE-Chirp^®^ LS spectra were 1.6-2.5x wider than those of the 2-1-2 tones at 100 dB pe SPL (-3 dB, Table 1). The exception to this was seen in the acoustic spectra for 4000-Hz NBchirps, where peak and geometric mean frequencies were lower and bandwidths less-wide (in relation to nominal frequency) than for lower frequency stimuli. As demonstrated below, this exception for 4000 Hz was due to the removal of higher frequencies by the earphone and acoustic coupler; this exception disappeared in the electrical spectra.

Table 1 shows that the -20-dB bandwidth of the NB CE-Chirp^®^ LS spectra at 500, 1000, 2000, and 4000 Hz were also considerably wider than those of the 2-1-2 tones. The -20-dB bandwidths showed that at 500 Hz, the NB CE-Chirp^®^ LS spectrum bandwidth was nearly twice as wide (1.8x) compared to that of the 2-1-2 tone (at 100 dB pe SPL). Similarly, the 1000-Hz and 2000-Hz NB CE-Chirp^®^ LS spectra were 1.8 and 1.9 times wider at 100 dB pe SPL compared to those of the 2-1-2 tones (-20 dB, Table 1). A slightly smaller difference, 1.4 times wider, was found when comparing NB CE-Chirp^®^ LS to 2-1-2 tone spectra for 4000 Hz (-20 dB, Table 1).

Figure 2 compares the electric and acoustic spectra for 500-, 1000-, 2000-, and 4000-Hz NB CE-Chirp^®^ LS and 2-1-2 stimuli. Table 2 compares measures of the electric spectra for NB CE-Chirp^®^ LS versus those of 2-1-2 tones at 100 dB pe SPL. Generally, the bandwidth for all the NB CE-Chirp^®^ LS electric spectra at -3 dB was approximately 1 octave; the -3-dB bandwidth for the 2-1-2 tones was 0.4 octave. The -3-dB bandwidths obtained for the 2-1-2 tones showed no clear differences between the acoustic and electric spectra (Table 1 versus Table 2). This was not the case for the NB CE-Chirp^®^ LS stimuli: the -3-dB bandwidths of the 1000- and 4000-Hz NB CE-Chirp^®^ LS electric spectra were about 0.4 - 0.5 octaves larger compared to their acoustic spectra (no differences were seen in the -3-dB bandwidths between the electric and acoustic spectra of the 500 and 2000 Hz NB CE-Chirp^®^ LS stimuli) (Table 1 versus 2). The differences for the 1000-Hz NB CE-Chirp^®^ stimuli appeared as a small dip in the acoustic spectra just below 1000 Hz, not present in the electric spectra. The differences for the 4000-Hz NB CE-Chirp^®^ stimuli appeared as a significant reduction in energy above 4000 Hz in the acoustic spectra, not seen in the electric spectra. As noted above, the reduction above 4000 Hz was due to the ER-3A insert earphone rolloff above this frequency.

**Figure 2.**
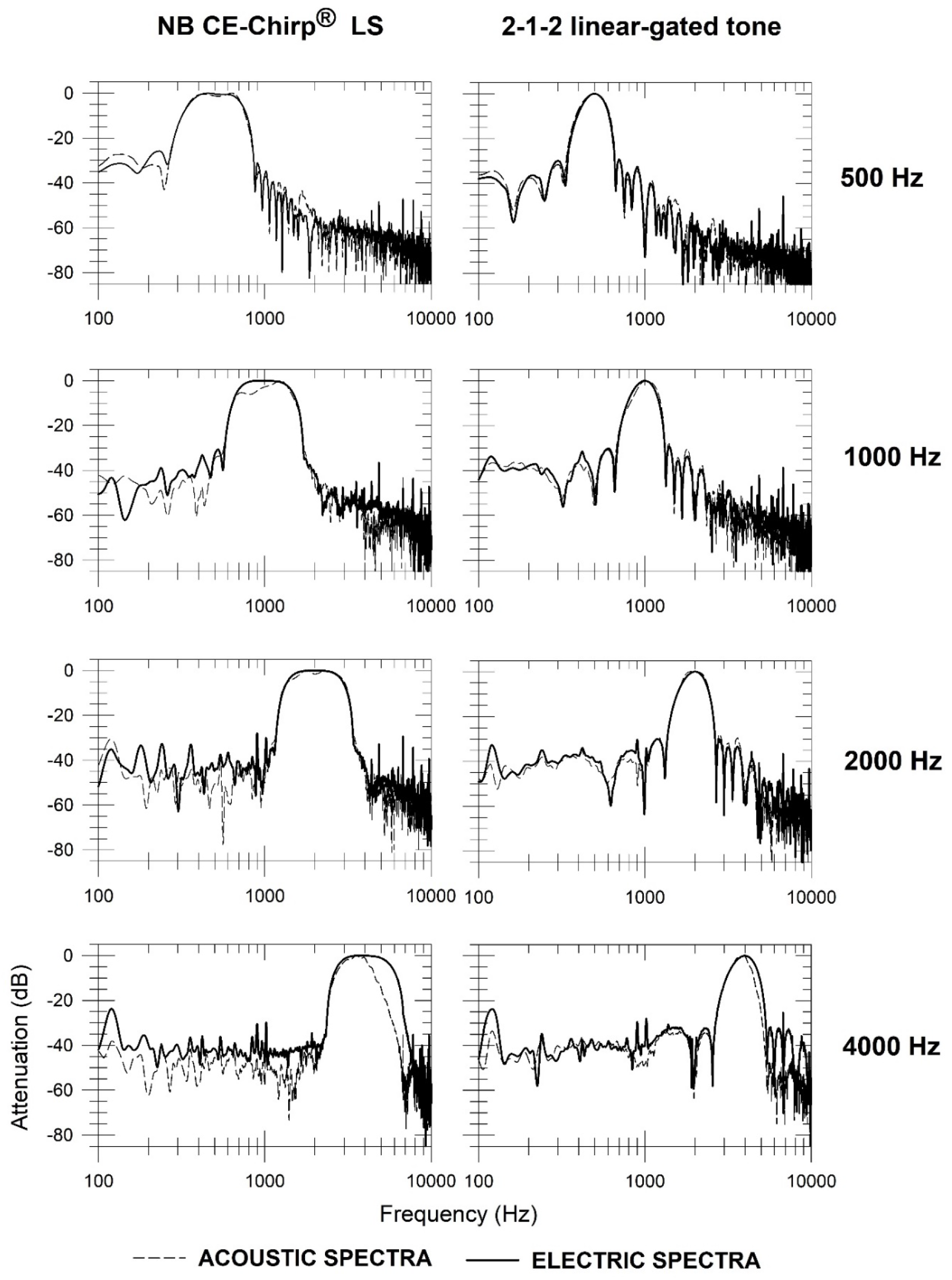
Electric and acoustic spectra for NB CE-Chirp® LS (left) and 2-1-2 (right) stimuli for 500, 1000, 2000, and 4000 Hz. (Acoustic spectra: dashed lines; Electric spectra: solid lines.)

**Table 2.** Electric spectra for NB CE-Chirp^®^ LS stimuli versus 2-1-2 tones at 100 dB pe SPL.

| Nominal<br>frequency (Hz) | Stimulus type | Peak Frequency (Hz) | CF (Geometric mean, Hz) |  | Bandwidth (Hz) |  | Bandwidth (octave) |  |
| --- | --- | --- | --- | --- | --- | --- | --- | --- |
|  |  |  | -3dB | -20dB | -3dB | -20dB | -3dB | -20dB |
| 500 | NB CE-Chirp<sup>®</sup> LS | 452 | 506 | 488 | 324 | 549 | 0.9 | 1.6 |
|  | 2-1-2 tone | 495 | 496 | 478 | 125 | 284 | 0.4 | 0.9 |
| 1000 | NB CE-Chirp<sup>®</sup> LS | 883 | 998 | 994 | 684 | 1067 | 1.0 | 1.5 |
|  | 2-1-2 tone | 1001 | 992 | 959 | 250 | 566 | 0.4 | 0.9 |
| 2000 | NB CE-Chirp<sup>®</sup> LS | 2078 | 2003 | 1997 | 1395 | 2130 | 1.0 | 1.5 |
|  | 2-1-2 tone | 2003 | 1986 | 1916 | 505 | 1144 | 0.4 | 0.9 |
| 4000 | NB CE-Chirp<sup>®</sup> LS | 3779 | 3952 | 4015 | 2633 | 4292 | 0.9 | 1.5 |
|  | 2-1-2 tone | 3994 | 3957 | 3809 | 1038 | 2371 | 0.4 | 0.9 |

Peak Frequency: Frequency with highest amplitude; GM: Geometric mean; BW: Bandwidth; NBCh: NB CE-Chirp<sup>®</sup> LS; 212 TB: 2-1-2 tone; -20dB: measured at 20 dB below maximum amplitude; -3dB: measured at 3 dB below maximum amplitude

The measurements at the -20 dB point shown for 100 dB pe SPL stimuli in Tables 1 versus 2 demonstrated that there were no large differences in bandwidth between acoustic and electric spectra for the 2-1-2 tone or NB CE-Chirp^®^ LS stimuli, except for 4000 Hz. For 4000-Hz 2-1-2 tones, the width of the acoustic spectra at -20 dB was similar to that of the electric counterpart; the 326 Hz difference is equivalent to only 0.1 octaves. In contrast, the spectra for the NB CE-Chirp^®^ LS stimuli showed that the 4000-Hz acoustic spectra bandwidth (at -20 dB) was about 1423 Hz (equivalent to 0.4 octaves) narrower than that of the electric spectra (Figure 2 and rTables 1 & 2). Again, this is mainly because the frequency response characteristics of the ER-3A earphones caused a considerable reduction in the higher frequency energy (above 4000 Hz), which affects the wider 4000-Hz NB CE-Chirp^®^ LS acoustic spectra more than the 2-1-2 spectra.

Using either the -3-dB or the -20-dB measures, the bandwidths for the NB CE-Chirp^®^ LS stimuli were much wider than those for the 2-1-2 tones (1.6-2.5 times wider). Overall, the spectra shown in Figures 1 and 2 indicated the NB CE-Chirp^®^ LS stimuli contained energy over a wider range than the 2-1-2 tones. However, the differences between the stimuli might not be fully accounted for by the above bandwidth measures. Thus, in order to assess the relative contributions of frequencies near the stimulus centre frequency versus those from frequencies further away, the acoustic waveforms (and electric waveforms for the 4000-Hz stimuli) were stopband filtered using Sigview software. As shown in Table 3, the narrow 0.5- and 1-octave-wide stopband filters had a greater effect on the 2-1-2 tones compared to the NB CE-Chirp^®^ LS stimuli, indicating most of the 2-1-2 tones’ energy was within ±0.5 octave of the centre frequency -- only 3% to 5% of the unfiltered energy remained after filtering with a 1-octave-wide stopband filter. In contrast, even after filtering with a 1-octave-wide stopband, 30% to 41% of the unfiltered energy remained for the NB CE-Chirp^®^ LS stimuli (excluding the 4000-Hz acoustic spectra) (Table 3). The narrow 0.5- and 1-octave-wide stopband filters had similar effects on the 4000-Hz NB CE-Chirp^®^ LS and 2-1-2 *acoustic* spectra. This is because the frequency response characteristics of the ER-3A earphones caused a considerable reduction in the higher frequency energy of the 4000-Hz NB CE-Chirp^®^ LS acoustic spectra, more than that of the 2-1-2 spectra. In summary, the results shown in Table 3 indicated the 2-1-2 tones concentrated their energy to within ±0.5 octaves of their CF, whereas the energy of the NB CE-Chirp^®^ LS stimuli was spread out over a wider area to within approximately ±0.75 octave of stimulus centre frequency.

**Table 3.**
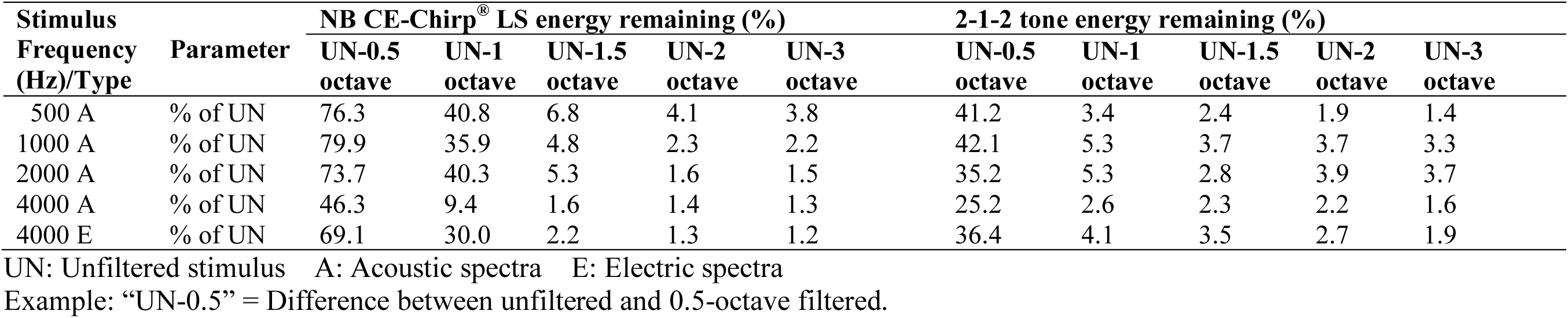
Percentage of unfiltered energy remaining after 0.5-, 1-, 1.5-, 2-, and 3-octave-wide stopband filtering, centered on stimulus geometric mean frequency (measured at -20 dB).

## Discussion

The largest differences between the spectra for NB CE-Chirp^®^ LS stimuli and 2-1-2 tones were seen for the bandwidths (whether measured -3 dB or -20 dB), which showed that, for all tested frequencies, the spectra for the NB CE-Chirp^®^ LS stimuli were considerably wider (1.6x to 2.5x) than those of the 2-1-2 tones. Both electric and acoustic spectra showed the NB CE-Chirp^®^ LS stimuli were considerably wider than those for 2-1-2 tones. The difference between the acoustic versus electric spectra of the 4000-Hz NB CE-Chirp^®^ LS is due to the frequency response of the ER-3A earphones, which reduce energy above 4000 Hz. The acoustic spectra of the 4000-Hz 2-1-2 tones also small showed reductions compared to their electric spectra energy, but this did not affect the bandwidth measure (affecting only the upper side lobes below -20 dB). In practical terms, if one is using earphones such as ER-3A insert phones, the bandwidth of the

NB CE-Chirp^®^ LS at 4000 Hz will not be as wide (in octaves) as for lower frequencies. However, if using earphones with a wider frequency response (e.g., ER-1 or ER-2), then the 4000-Hz NB CE-Chirp^®^ LS acoustic spectra will likely be as wide as the lower frequency NB CE-Chirp^®^ LS stimuli.

The stopband filter analysis results were consistent with the above spectral bandwidth measures, indicating that when one octave around the centre frequency was stopband filtered, 30-40% of the energy remained for the NB CE-Chirp^®^ LS stimulus, in contrast to only 3-5% for the 2-1-2 tones.

Elberling and colleagues used the -3 dB cut-off point to describe the amplitude spectra of NB CE-Chirp^®^ stimuli, indicating their adherence to the characteristics of a conventional one-octave-band filter (Elberling & Don, 2010; Gøtsche-Rasmussen et al., 2012). Additionally, the frequency response characteristics of the earphone and acoustic coupler shape the acoustic spectra (Elberling & Don, 2010; Gøtsche-Rasmussen et al., 2012). The description above agrees well with the -3-dB bandwidths obtained in the current study, but it is important to note that the -3-dB bandwidth underestimates the effective bandwidth that contributes to a response. For example, several studies, including high-pass noise/derived response, notched noise, and pure-tone masking experiments, have shown that the ABR to tonal stimuli includes contributions from frequencies in the stimuli much beyond the -3 dB cut-off point (even more so when hearing thresholds differ across frequencies) (e.g., Oates & Stapells, 1997; Purdy & Abbas, 2002). Thus, the -20-dB bandwidth is a more realistic/appropriate estimate of physiologically effective bandwidth contributing to a response.

The wider bandwidths of NB CE-Chirp^®^ LS stimuli compared to 2-1-2 tone stimuli bring up a long-standing question in audiometry in general and ABR audiometry, specifically “What is the acceptable bandwidth for audiometric stimuli (in general) and for evoked potential stimuli in particular?” In behavioural audiometry, pure-tone stimuli, effectively composed of a single frequency, are the standard stimulus. For testing in soundfield and/or testing of infants and young children, warble tones are often used (ANSI S3.6, 2018); again, stimuli of very narrow bandwidth. In contrast, 1/3- to 1/2-octave-wide “narrowband noise” is considered too wide for measuring the behavioural audiogram (ANSI S3.6, 2018). The acoustic frequency specificity of stimuli used to elicit the ABR has long been a topic of much discussion (and debate). The bandwidths of tonal stimuli used to elicit the ABR are necessarily wider, as relatively shorter stimuli are required (e.g., Stapells & Picton, 1981). Davis and colleagues (Davis, 1976; Davis et al., 1984) concluded that the 2-1-2 cycle tone provides the best trade-off between frequency-specificity and ABR amplitude. Many studies have since shown reasonably accurate estimation of pure-tone behavioural thresholds by the ABR to 2-1-2 (or similar) tones in individuals with hearing loss (for review, see Small & Stapells, 2017).

In spite of evidence in support of 2-1-2 stimuli, some researchers have suggested the ABRs to 2-1-2 tones are not sufficiently frequency specific (e.g., Gorga & Thornton, 1989; Laukli, 2014). For example, there were suggestions that ABRs to low-frequency 2-1-2 (and similar) stimuli are mediated primarily by contributions from the basal portion of the cochlea (Burkard & Hecox, 1983; Laukli et al., 1988). Some investigators argued that the spectral differences observed between linear versus nonlinear gating functions (e.g., the reduced sidelobe energy of the Blackman-gated tone when compared to 2-1-2 tones) should improve the frequency specificity of the ABR (Gorga & Thornton, 1989). Subsequent studies, however, demonstrated no effective difference in the frequency specificity of the ABRs to these stimuli (Oates & Stapells, 1997; Purdy & Abbas, 2002).

Although the clinical evidence base for NBchirps is growing, there are very few studies comparing NBchirp to 2-1-2 ABR amplitudes and/or thresholds in individuals with hearing loss (Talaat et al., 2019) and, importantly, no study has assessed these in individuals with steeply sloping hearing loss. The advantages/disadvantages of ABRs to NBchirps versus 2-1-2 tones when assessing infants with hearing loss are thus not yet known. As demonstrated by the present study, NBchirps show even wider frequency spectra than those for 2-1-2 tones. The 1.6x to 2.5x wider bandwidth of NB CE-Chirp^®^ LS stimuli compared to 2-1-2 tones (e.g., 2000-Hz NBchirps bandwidth at -20 dB was 2130 Hz, compared to 1144 Hz for 2-1-2 tones; Table 2) should result in even greater problems with frequency specificity when assessing individuals with hearing loss, and certainly require investigation and deserve discussion. It is possible, however, that the relatively large spectral differences between 2-1-2 tones and NB CE-Chirp^®^ LS stimuli may not, in practice, translate into large differences in the cochlear place specificity of ABRs to these two stimuli (similar to the results comparing ABRs to linear 2-1-2 versus 5-cycle Blackman tones).

Future research with individuals with steep hearing loss, as well as masking studies (e.g., high-pass noise) should compare the cochlear place specificity (i.e., the effective spectra) of NB CE-Chirp^®^ LS and 2-1-2 tones.

In conclusion, NB CE-Chirp^®^ LS stimuli have poorer frequency (acoustic) specificity compared to the 2-1-2 tones that have been used for many years to assess infant ABR thresholds. As reported by several studies, the ABRs to NBchirps are larger than those to 2-1-2 tones in certain stimulus conditions (Bell et al., 2002; Cobb & Stuart, 2016; Dzulkarnain et al., 2018; Ferm & Lightfoot, 2015; Rodrigues et al., 2013). Larger response amplitudes result in better signal-to-noise ratios and faster testing. It has been suggested that these larger amplitudes are due to the optimized timing of frequency components within the NBchirp (e.g., Elberling & Don, 2010). However, the larger amplitudes might be more simply explained by the NBchirp’s broader spectrum (e.g., Bell et al., 2002; Cobb & Stuart, 2016), as stimuli with wider spectra produce ABRs with larger amplitudes. Further research focussing on the cochlear place specificity of the ABR to NB CE-Chirp^®^ LS stimuli is required.

## Ethical approval

Not required. No testing of, nor data from, humans or animals.

## Data availability

The stimulus waveforms recorded and analyzed in the current study are available from the corresponding author on reasonable request.

## Acknowledgements

This study was carried-out in Dr. Susan Small’s laboratory under her initial supervision. Sadly, Dr. Small died before completion of this study. The authors thank Bue Kristensen at Interacoustics and for providing the Eclipse ABR system for presentation of the stimuli.

## Footnotes

1 Using Fedtke and Richter’s (2007) RETSPL values, after correction for the use of different couplers (IEC 60126 2-cc vs IEC 60711; Haughton, 2006), 60 dB pe SPL corresponds to 41, 45, 38 and 37 dB nHL for 500-, 1000-, 2000- and 4000-Hz 2-1-2 cycle stimuli, respectively. NB CE-Chirp LS stimuli presented at 60 dB pe SPL (after correction for use of 2-cc coupler) correspond to 40, 41, 37, and 36 dB nHL for 500, 1000, 2000, and 4000 Hz, respectively (Gøtsche-Rasmussen et al., 2012).

2 The timing of NB CE-Chirp^®^ LS stimuli is typically adjusted in an attempt to result in similar wave V latencies for different stimulus frequencies. These onset timing differences are not reflected in the waveforms of Figure 1.

3 Confirmed by email communication with Interacoustics

